# Perceived usability and usefulness of a clinical decision-support application among newly graduated physicians in rural areas: a mixed-methods study

**DOI:** 10.64898/2026.08.18.26360759

**Authors:** Kelly De la Cruz-Torralva, Paquita Díaz-Sánchez, Stefan Escobar-Agreda, Leonardo Rojas-Mezarina

**Affiliations:** Unidad de Telesalud, Facultad de Medicina, Universidad Nacional Mayor de San Marcos, Lima, Perú; Unidad de Posgrado, Universidad San Ignacio de Loyola, Lima, Perú

**Author notes:** **Corresponding author:** Stefan Escobar-Agreda, Address: 550 La Fontana Avenue, La Molina, Lima, Peru.

**Keywords:** Digital health, Clinical decision support, Mobile health, Rural health, Usability

## Abstract

Mobile clinical-support applications can facilitate access to evidence-based information at the point of care, but evidence on their usability and perceived usefulness among newly graduated physicians working in health facilities with limited capacity is scarce. We assessed physicians’ experiences with BMJ Best Practice using a convergent mixed-methods study. All 81 eligible physicians assigned to rural facilities were invited; 32 enrolled and received application access and training. After three months, participants completed an online survey, and 23 reported using the application. Ten physicians reporting the highest consultation frequency were purposively selected for semi-structured interviews. Survey findings showed a predominantly favorable perception of usability: for most items, 70%–90% of participants agreed or strongly agreed with the statements assessed. Among users, 14 of 23 (60.9%) used the mobile application and 9 (39.1%) used the web version. Interviews indicated that participants valued rapid searches, organized and evidence-based information, and support for diagnostic reasoning, referral decisions, learning, and clinical confidence. Barriers included limited connectivity, difficulties searching in Spanish, automatic updates, challenges locating or using some calculators, and treatment information that was sometimes insufficiently specific. Most importantly, participants could not always implement recommendations because suggested medicines, diagnostic tests, or other resources were unavailable in their facilities. Mobile clinical-support applications may complement decision-making and learning among early-career physicians in rural primary care. However, their practical value depends not only on usability and evidence quality, but also on adaptation to users’ language, workflow, connectivity, and local service capacity.

**AUTHOR SUMMARY:** We evaluated whether BMJ Best Practice, a mobile application that provides evidence-based clinical information, was useful to newly graduated physicians completing SERUMS in rural Peru. Thirty-two physicians had access to the application for three months; 23 reported using it and 10 participated in interviews. Users valued its speed, information organization, and support for diagnostic reasoning, referral decisions, and greater confidence during patient care. However, access to a global tool did not resolve constraints in the practice environment. Connectivity, searches in Spanish, automatic updates, and the performance of some calculators hindered use. Moreover, several recommendations required medicines, diagnostic tests, or other resources that were unavailable in rural primary-care facilities. Our findings suggest that these applications may support learning and decision-making among early-career physicians, but their usefulness depends on adaptation to local language, workflow, and service capacity. Studies using objective usage metrics and clinical outcomes are also needed.

## INTRODUCTION

Mobile clinical-support applications provide point-of-care access to evidence-based information, clinical guidelines, algorithms, and tools such as medical calculators. These tools have been described as resources for supporting clinical decision-making and providing timely access to clinical information during patient care (1–3). They are frequently used by medical students, physicians in training, and early-career physicians for educational support and immediate access to information in clinical practice (3,4).

In Peru, the Rural and Urban-Marginal Health Service (SERUMS) is a one-year program that places health professionals in rural and urban-marginal communities; completing the program is a requirement for professionals seeking to enter the public health system. Physicians completing SERUMS commonly provide outpatient care with considerable autonomy in primary-care facilities with heterogeneous service capacity. In these settings, decision-making may be constrained by limited clinical experience, restricted access to diagnostic tests, supplies and equipment, and limited point-of-care specialist support (5). Clinical-support applications could complement other support strategies in this context, such as remote mentoring, for which barriers related to connectivity, confidence in formulating questions, and the need for timely answers during clinical practice have been described (6).

Some clinical-support applications combine evidence-based information, clinical tools, and offline-access options, potentially facilitating their use where connectivity or specialist support is limited. However, evidence on the usability and perceived usefulness of these tools among newly graduated physicians completing SERUMS remains limited. This study therefore aimed to evaluate the perceived usability and usefulness of a mobile clinical-support application among newly graduated physicians during SERUMS in Peru.

## METHODS

### Study design

We conducted a mixed-methods study with convergent integration. The quantitative component described self-reported application use and perceived usability and usefulness through an online survey. The qualitative component used semi-structured interviews to explore perceptions of application-feature usability and its usefulness in supporting clinical decision-making during SERUMS. Both components were planned at study inception and analyzed separately; the qualitative subgroup was selected after the survey, and findings were integrated during interpretation and preparation of the Discussion. Reporting was guided by the Good Reporting of a Mixed Methods Study (GRAMMS) recommendations for mixed-methods health-services research (7).

### Study population and sampling

Eligible participants were newly graduated physicians from the National University of San Marcos who completed SERUMS in Peru during 2023–2024 and were assigned to rural health facilities. None of the participants worked in an urban-marginal facility. Because the number of eligible physicians was limited, all 81 eligible physicians were invited to participate; no sampling was applied for the quantitative component.

For the qualitative component, we purposively sampled participants who reported the highest frequency of consultations in the application in the quantitative survey. This strategy was intended to elicit richer accounts of the user experience. The qualitative component was not designed to explain specific quantitative results, but to provide a deeper understanding of perceptions of use, usability, and clinical usefulness. Interviews continued until thematic saturation, defined as the point at which additional interviews contributed no substantially new information relevant to the study objectives. The decision to stop interviewing also considered the adequacy of the information obtained, the specificity of the objectives, and the quality of the interview dialogue (8).

### Clinical-support application

We used BMJ Best Practice, a clinical-support application developed by BMJ Publishing Group that provides evidence-based clinical information, treatment guidance, diagnostic tools, and educational materials (9). The application is designed to support point-of-care clinical decision-making, particularly where timely access to clinical information may be limited. During the study, the interface and most content were available in English, although a limited set of topics could be accessed in Spanish.

### Enrollment, access, and training

Enrollment took place between May and July 2024. The university provided contact information for the 81 eligible newly graduated physicians completing SERUMS in 2023–2024, and all 81 were contacted by email. The email included a link to an online form that first presented information about the study purpose, voluntary participation, data confidentiality, and electronic informed consent. Thirty-two physicians completed the form, and all 32 provided electronic consent; no individual who completed the form declined consent. Enrolled participants received access credentials and initial training, followed by three months of access to the application. Each participant received a username and password that provided three months of application access from the date of enrollment. Use of a mobile device with an up-to-date operating system (iOS ≥10.0 or Android ≥7.0), a compatible browser (Chrome, Firefox, or Safari), and a stable internet connection (≥3G or Wi-Fi) was recommended. Participants received synchronous virtual training, supplemented by audiovisual materials covering download, navigation, personalization, general use of the application, and access to continuing-learning certificates. A contact telephone number was also provided for questions about access to and use of the application during the study period.

### Quantitative component: variables and instrument

At the end of the three-month access period, an online survey was distributed through REDCap. The survey collected participant characteristics (age and sex), characteristics of the rural health facility where they completed SERUMS (facility level and region), and self-reported application-use characteristics, including frequency of use, number of consultations, and most-used features. The study did not have access to objective usage logs or internal application analytics.

Perceived usability was assessed using a 17-item Spanish version based on the adaptation of the Computer System Usability Questionnaire (CSUQ) used by English et al. to evaluate the usability of a mobile health application among health workers in a resource-limited setting (10). Items were translated and adapted to the study context and assessed ease of use, speed, comfort, recovery from errors, clarity of information, on-screen organization, overall satisfaction, and availability of expected functions. Responses used a five-point Likert scale ranging from ‘strongly disagree’ to ‘strongly agree.’ Because this adapted version did not undergo formal psychometric validation, findings were interpreted descriptively as self-reported perceptions of usability.

### Qualitative component: interviews

Individual semi-structured interviews were conducted in Spanish by video call. The interview guide was developed from the study objectives and explored participants’ perceptions of the usability of application features, barriers and facilitators to use, and usefulness in supporting clinical decision-making in rural settings. Two researchers with previous experience in qualitative and mixed-methods digital-health research conducted the interviews. Each interview lasted approximately 20 minutes and was audio-recorded with participants’ prior informed consent.

### Quantitative analysis

The quantitative analysis was descriptive and was performed in Stata 17. Categorical variables were summarized using absolute frequencies and percentages. Numerical variables were summarized using means and standard deviations or medians and interquartile ranges, according to their distribution. Tables described participant characteristics, and figures presented self-reported application-use patterns and perceived user experience.

### Qualitative analysis

The qualitative analysis followed Braun and Clarke’s thematic-analysis approach (11). Two researchers transcribed the interviews verbatim and checked the transcripts against the audio recordings. One researcher performed initial coding in ATLAS.ti 9, guided by the study objectives while remaining open to emergent codes related to usability, clinical usefulness, learning, barriers to use, and recommendations for improvement. Codes were then iteratively grouped into provisional themes. Two researchers reviewed the themes for internal coherence, conceptual distinctiveness, and relevance to the study objectives. Disagreements were resolved through discussion until consensus was reached. Final themes were operationally defined and documented in a codebook with examples from the transcripts. Representative quotations were selected and related to the study objectives. For this English-language manuscript, quotations originally given in Spanish were translated into English while preserving their meaning and conversational tone.

### Mixed-methods integration

The quantitative and qualitative components were integrated during interpretation of the findings and preparation of the Discussion. Descriptive survey findings were compared with themes emerging from the interviews to identify convergence, divergence, and complementary explanations regarding self-reported use, perceived usability, clinical usefulness, barriers to use, and recommendations for improving the application.

### Ethical considerations

The study was approved by the Research Ethics Committee of the Faculty of Medicine of the National University of San Marcos (approval number 0293-2023). All participants provided electronic informed consent before participation. Additional consent for audio recording was obtained for the interviews. No financial compensation was offered. Data were anonymized for analysis and handled in accordance with confidentiality and information-protection principles and the ethical principles of the Declaration of Helsinki.

## RESULTS

### Quantitative results

Of the 81 eligible physicians contacted, 32 completed the online form and provided electronic consent (Table 1). All 32 worked in rural health facilities. Their mean age was 28.0 ± 2.3 years, and 18 (56.3%) were women. Regarding English proficiency, 13 (40.6%) reported an intermediate level and 14 (43.8%) an advanced level. Most worked in level I-2 health facilities (22; 68.8%), and 15 (46.9%) were located in the Central macro-region. Fifteen participants (46.9%) accessed BMJ Best Practice once or twice per week, whereas 9 (28.1%) reported not using it during the evaluation period.

**Table 1.** Characteristics of participating physicians (N = 32)

|  | n | (%) |
| --- | --- | --- |
| Total | 32 | (100.0) |
| Age, mean ± SD | 28.0 ± 2.3 | |
| Sex |  |  |
| Female | 18 | (56.3) |
| Male | 14 | (43.8) |
| English proficiency |  |  |
| Basic | 5 | (15.6) |
| Intermediate | 13 | (40.6) |
| Advanced | 14 | (43.8) |
| Health-facility level |  |  |
| I-1 | 5 | (15.6) |
| I-2 | 22 | (68.8) |
| I-3 | 3 | (9.4) |
| I-4 | 2 | (6.3) |
| Macro-region |  |  |
| Central | 15 | (46.9) |
| Lima–Callao | 3 | (9.4) |
| Northern | 7 | (21.9) |
| Eastern | 3 | (9.4) |
| Southern | 4 | (12.5) |
| Frequency of use |  |  |
| Did not use | 9 | (28.1) |
| Once or twice per week | 15 | (46.9) |
| Three times per week | 5 | (15.6) |
| Four or five times per week | 3 | (9.4) |

Among physicians who reported using the application (n = 23), 13 (56.5%) consulted BMJ Best Practice after the patient encounter, 6 (26.1%) during the consultation while visible to the patient, and 4 (17.4%) during the consultation without the patient observing them. Regarding platform, 14 participants (60.9%) used the mobile application and 9 (39.1%) used the web version. Fifteen of the 23 users (65.2%) explored all three principal modules available: clinical updates, webinars, and searches for signs, symptoms, or diseases. Other resources, such as calculators, videos, patient leaflets, and podcasts, were used infrequently or not at all (Fig 1).

**Fig 1.**
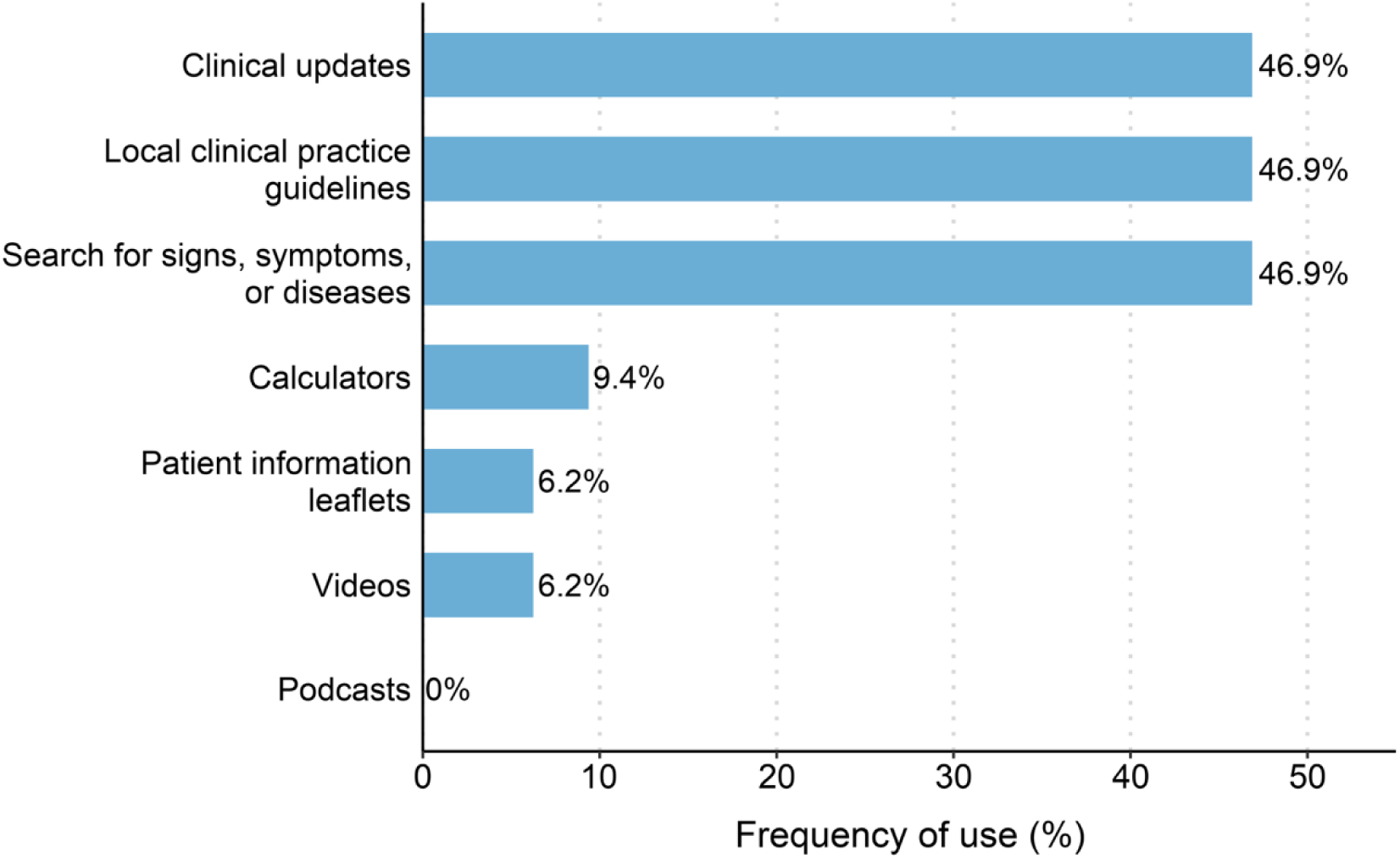
Use of BMJ Best Practice modules among physicians completing SERUMS (N = 23).

The perceived-usability assessment is presented in Fig 2. For most items, between 70% and 90% of participants agreed or strongly agreed with the statements assessed. Overall, the findings suggest a predominantly positive perception of application usability.

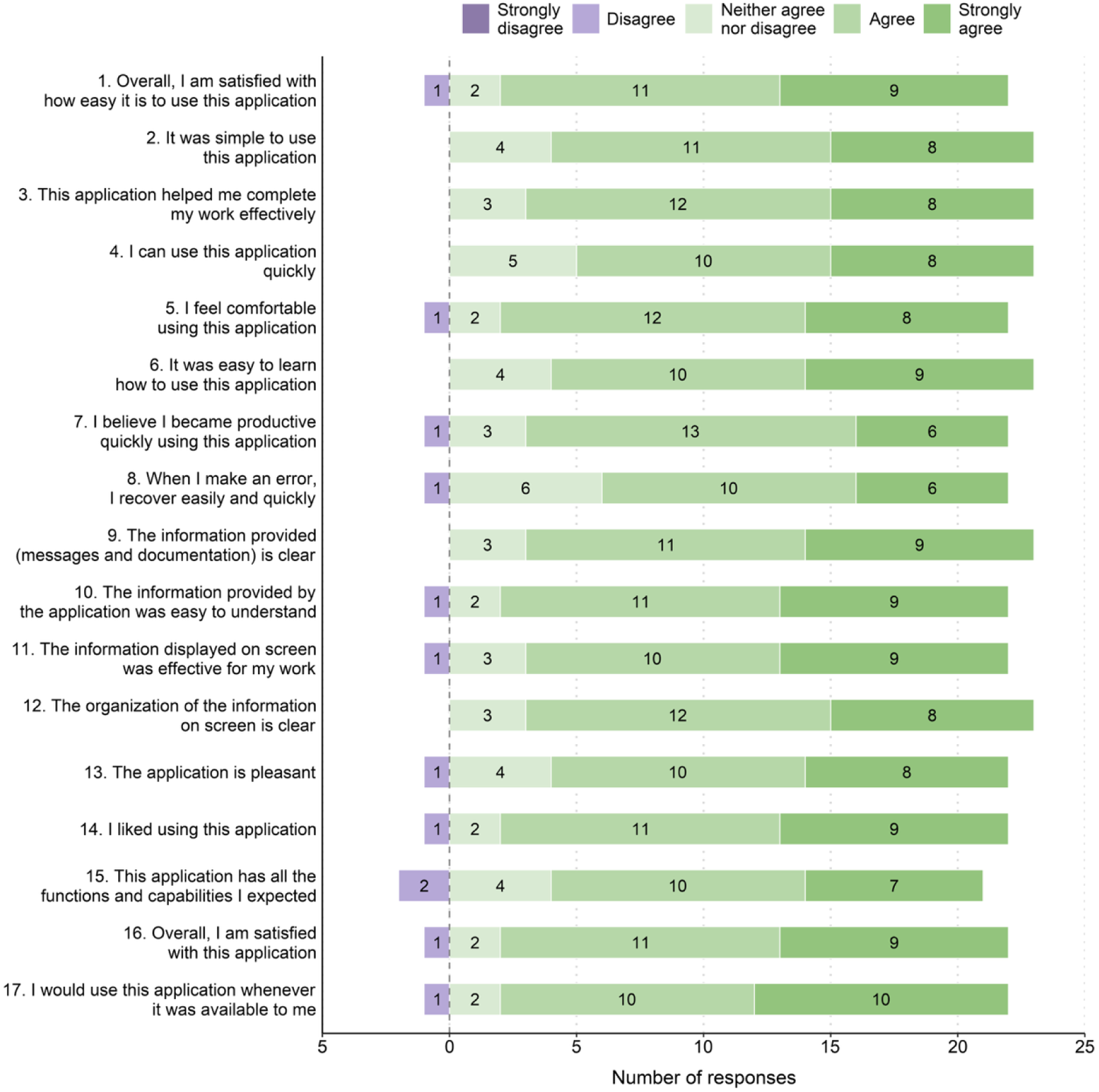

### Qualitative results

#### Application usability

Participants valued the speed of the application’s search function, particularly because it facilitated access to clinical information during patient care. However, they also described initial difficulty finding specific information, mainly because they had to refine search terms or use synonyms in English. Although these difficulties tended to lessen with continued use, limitations related to searching in Spanish persisted and sometimes produced inadequate results.

> ‘There were some terms that did not appear when I searched for them, so I had to search in another way (…) or search in English (…) you had to use certain words to find some things.’ (Participant 2)

Clinical tools such as calculators were considered potentially useful for complementing decision-making. However, some participants reported difficulty locating certain scores or specific tools, which sometimes limited their use.

> ‘With the calculators, when I wanted to use a score or something like that, I did find it difficult because I could not find the score I wanted. Sometimes I had to search for terms similar to what I was looking for to find it in the application.’ (Participant 1)

The application’s content was valued for its clarity, organization, and visual presentation. Participants highlighted its orderly item structure, on-screen display, and availability of multimedia resources such as videos, images, and translation options. These elements were perceived as helpful for understanding information, identifying clinical signs, and guiding diagnostic reasoning. The management section was also considered useful because its recommendations were clear and structured.

> ‘The items are organized within the application (…), even the audio for playing the videos is quite good, and it also lets you use a translator. So I had no problems, and using the application was quite useful.’ (Participant 1)

Regarding technical performance, participants preferred the mobile version because it was practical, quick to access, and intuitive, which facilitated use during the working day. In contrast, the web version was considered less convenient, particularly where connectivity was limited, although it was used as an alternative when the application could not be installed.

> ‘I use my phone more because I always have it at hand, and the application is much more pleasant (…) if you needed to look something up (…) it was much more practical to do it on the phone (…) than on a desktop computer.’ (Participant 1)

Some users also reported initial difficulty becoming familiar with the interface, as well as limitations related to having to enter multiple sections repeatedly and the absence of quick summaries. These limitations reduced perceived efficiency compared with other applications.

> ‘During the first few days it was a little difficult because I did not know the application very well, but as the days and weeks went by, I did not have any other problems.’ (Participant 5)

#### Usefulness in clinical practice and learning

Participants stated that the application supported diagnosis and clinical decision-making, mainly through searches for diseases based on signs and symptoms, clinical algorithms, and criteria for referral or transfer to facilities with greater service capacity. These features were perceived as useful for distinguishing cases that could be managed in primary care from those requiring referral, especially in complex or potentially urgent clinical situations.

> ‘Yes, it has definitely helped me. Once, a patient came to my health post with symptoms of diabetic ketoacidosis. I was able to check the application very quickly at that moment; it provided good algorithms. It was during a health campaign, so being able to consult up-to-date sources right then helped me make decisions about referring the patient. Based on that information, I could also request the ambulance so that it would arrive more quickly.’ (Participant 10)

Nevertheless, participants identified limitations in the therapeutic component. They reported that the information could be general and, in some cases, did not provide exact doses or sufficiently locally applicable instructions. Some physicians therefore supplemented the information with other sources, especially when seeking prescribing guidance.

> ‘If I had a question about a dose or, above all, about prescribing treatment, the application provided general information. It did not tell you exactly what to use in a particular case; it gave you guidance, and then directed you to other pages where you could find the information you needed. That was a limitation for me: treatment information was a little more difficult and not very quick to find. I necessarily had to seek help from other pages or related articles.’ (Participant 1)

The application was also valued for providing rapid access to up-to-date, evidence-based clinical information, which increased some participants’ confidence in decision-making. This support was particularly valued when there was little time to verify the quality of other sources.

> ‘When I could and I had a question, I checked it (…). It gives you confidence because the information has a sound basis. Sometimes you search online and it simply says, “give this,” and there is no time to verify whether the evidence is correct, appropriate, and current. But when you search BMJ, you are confident that it is at least the most up to date (…) and you feel completely confident.’ (Participant 4)

One of the main challenges was the gap between application recommendations and the resources actually available in rural facilities. Participants reported that some suggested diagnostic tests, treatments, or resources were unavailable in primary care, forcing them to adapt clinical decisions to local conditions. Some physicians also perceived the application as being oriented toward hospital settings with greater service capacity, limiting its full applicability during SERUMS.

> ‘Because of the range of conditions seen in a hospital (…) I think it would help them more, because they have more ways to apply the information: they have the medicines mentioned and the diagnostic support suggested by the application. It could even help with their academic activities. In contrast, during SERUMS, if you are fortunate, your facility holds academic activities and you can search for information that helps with those too; but when applying it to the reality of SERUMS, the service capacity of each facility may not allow you to take full advantage of the application.’ (Participant 5)

Despite these limitations, participants valued the application as a source of support during the first months of rural service. Some used it both at the health facility and at home to reinforce their knowledge, review clinical uncertainties, and guide their practice.

#### Recommendations for application use

Participants suggested improvements to increase the application’s usefulness in rural clinical practice. First, they recommended including more images illustrating clinical signs so that users could compare visual findings and support diagnostic reasoning. They also suggested integrating ICD-10 codes into the platform because diagnostic coding is part of routine practice in primary care.

> ‘Add ICD-10 codes (…). At least in primary care, we always have to record everything correctly (…) and searching for them online is complicated.’ (Participant 3)

Another prominent recommendation concerned optimization of the update system. Users suggested that the application display a notification before running automatic updates because updates could take time and interfere with use in rural areas with limited connectivity. They also emphasized the need to strengthen offline access so that content could be consulted even when internet access was unavailable.

> ‘It should be possible to turn off automatic updates when there is an internet connection and, instead, be asked whether you want to update at that moment. Updating takes a long time, and we cannot use the application while it is updating; sometimes that is exactly when we need it.’ (Participant 7)

Finally, given the specific characteristics of rural practice, participants proposed that local professionals review or adapt the information so that clinical recommendations would better reflect the medicines, diagnostic tests, and other resources actually available in Peruvian primary care.

> ‘It would be good if Peruvian physicians could review it and tailor it more to our setting, particularly regarding medicines.’ (Participant 6)

## DISCUSSION

Among physicians who used BMJ Best Practice during SERUMS in Peru, the application was perceived as useful and favorably usable. Quantitative findings showed positive ratings for ease of use, clarity of information, content organization, and overall satisfaction. The interviews showed that these ratings were related to search speed, access to up-to-date clinical information, and perceived support for diagnostic reasoning and decisions to refer or transfer patients to facilities with greater service capacity. However, this favorable perception coexisted with technical, linguistic, and contextual barriers that limited use and applicability in rural primary-care facilities, including limited connectivity, difficulty searching in Spanish, the need to use English terms, automatic updates, problems with some calculators, and a mismatch between application recommendations and locally available resources.

The preference for the mobile application over the web version is consistent with literature describing frequent use of smartphones and mobile applications by physicians for medical education, clinical-information searches, decision support, and review of drug compendia (3). In our study, this preference was attributed to the practicality of the mobile phone, rapid access, and the ability to consult information during the working day. This finding is especially relevant for early-career physicians, for whom mobile technologies may serve as workplace-learning resources and provide support during the transition to more autonomous clinical practice (4,12). Nevertheless, almost one-third of participants reported not using the application during the evaluation period, suggesting that providing access, credentials, and initial training does not by itself ensure that a digital tool will be incorporated into clinical workflow. Future implementations should examine reasons for non-use in greater depth.

Regarding clinical usefulness, participants described the application as a rapid-reference resource that supported diagnostic reasoning, review of management options, and referral decisions. This finding is consistent with the role of point-of-care clinical-information resources, which are designed to provide health professionals with rapidly accessible, up-to-date summaries (13). It also aligns with studies of mobile applications and clinical decision-support systems in primary care, in which such tools have been evaluated as resources for organizing information, supporting clinical decisions, and facilitating the management of common conditions (1,2,14,15). However, BMJ Best Practice primarily functions as a point-of-care clinical-information resource rather than an automated system integrated into electronic health records or prescribing workflows. Therefore, literature on clinical decision support should be interpreted as contextual rather than as directly equivalent evidence for the tool evaluated here. Perceived usefulness was lower for specific therapeutic decisions, particularly when participants needed doses, locally available medicines, or recommendations applicable to primary care. The application was therefore valued more as a complementary source of information and guidance than as a sole source for prescribing or management decisions. A central tension emerged between the value of a global source of evidence-based clinical information and the need for recommendations that could be acted on locally. This tension is particularly relevant during SERUMS, when early-career physicians make decisions with limited access to specialist support, restricted diagnostic resources, and variable service capacity.

These findings should be interpreted cautiously in relation to potential clinical effects. Although clinical decision-support systems may improve care processes, effect sizes are variable and generally small to moderate (16). Evidence on mobile-device decision-support tools in primary care also remains limited and dependent on the implementation context (15). Because this study did not assess clinical outcomes, decision quality, medical errors, or patient outcomes, its findings represent evidence of perceived usability and usefulness rather than clinical effectiveness. Future studies should evaluate whether perceived gains in confidence and access to current information translate into objective improvements in clinical-decision quality, timeliness of referral, treatment appropriateness, or patient safety.

The gap between application recommendations and the resources available in rural primary-care facilities was a central finding. Participants reported that certain diagnostic tests, treatments, or other suggested resources were unavailable, limiting the direct applicability of the information. This finding is consistent with studies of young physicians in rural Peru that describe barriers involving infrastructure, supplies, equipment, and access to more experienced professionals (5). It also aligns with broader evidence on digital interventions in low- and middle-income countries, where sustainability depends on technical, organizational, human, and infrastructural factors, including connectivity, available resources, training, technical support, and adaptation to the local context (17,18). Thus, a mobile application may support decision-making but cannot by itself compensate for structural limitations in the health system (19).

The limited availability of medicines described by participants is also consistent with national indicators: in February 2024, at least 20% of health facilities in more than half of Peru’s regions had insufficient stocks or were out of stock of essential medicines (20). Although this indicator is not specific to SERUMS placements, it helps explain why a clinically valid treatment recommendation might not be actionable at a local facility and could require substitution, referral, or transfer.

The technical and usability barriers identified have practical implications for future implementation. Participants reported difficulty searching in Spanish, the need to use English terms, delays caused by automatic updates, problems with some calculators, and initial difficulty becoming familiar with the interface. These barriers are related to factors described in reviews of clinical decision-support adoption in primary care, where workflow integration, ease of use, relevance of recommendations, training, and user trust influence incorporation into clinical practice (21). Technical improvements should prioritize optimized Spanish-language search, offline access, configurable updates, and better performance of clinical tools such as calculators. From an implementation perspective, access should be accompanied by context-specific training, ongoing technical support, and guidance on use cases relevant to rural primary care.

The application’s language also warrants attention. Although a substantial proportion of participants reported intermediate or advanced English proficiency, the interviews showed that the need to search using English terms or synonyms could hinder rapid access to information. This barrier may have been attenuated by the language profile of this sample, but in broader implementation among physicians completing SERUMS it could affect usability, uptake, and equitable use among professionals with lower English proficiency. Linguistic localization should therefore extend beyond content translation to include adaptation to terminology used in everyday clinical practice, common primary-care diagnoses, ICD-10 codes, medicine availability, referral pathways, and visual resources relevant to rural care.

### Strengths and limitations

This study has several limitations. The sample was small, non-probabilistic, and limited to newly graduated physicians from one university, restricting the transferability of findings to other training contexts, regions, or cohorts of physicians completing SERUMS. Voluntary participation may also have favored physicians with greater interest in digital tools or evidence-based information. Nevertheless, all 81 eligible physicians were invited, and the study described the experience of the 32 physicians who completed the online form, provided electronic consent, and received access to the application.

Application use was self-reported, and objective usage logs and internal platform analytics were unavailable. Usability was likewise assessed through self-report rather than observation of performance on clinical tasks, and the Spanish-language instrument did not undergo formal psychometric validation. The qualitative component prioritized participants with the highest self-reported consultation frequency, providing more detailed information on user experience but potentially underrepresenting barriers among those who did not adopt the tool. Reasons for non-use were not specifically explored, even though almost one-third of participants did not use the application during the evaluation period. Finally, the three-month follow-up captured an initial experience but not sustained use or effects on knowledge, decision quality, medical errors, referrals, or patient outcomes.

Strengths include the use of a mixed-methods design in a real-world rural clinical setting. Combining a survey with interviews allowed us to describe patterns of use and perceived usability while also understanding barriers, facilitators, and contextual conditions relevant to adoption of a clinical-support application. The study also addresses an understudied population that is important to primary care in hard-to-reach areas. To our knowledge, it is among the first studies in Peru to evaluate the use experience of a mobile clinical-support application among newly graduated physicians during rural service.

### Conclusions and future research

Among physicians who used BMJ Best Practice during rural service in Peru, the application was perceived as useful and favorably usable. Its principal value was related to rapid access to current clinical information, support for diagnostic reasoning, and increased subjective confidence during decision-making. However, its use and benefit were constrained by technical, linguistic, and contextual barriers, particularly limited connectivity, difficulty searching in Spanish, automatic updates, and the gap between clinical recommendations and the resources available in rural primary-care facilities.

These findings suggest that mobile clinical-support applications may complement support strategies for early-career physicians but require local adaptation to be useful in rural settings. Future research should include larger and more diverse samples, explore reasons for non-use, incorporate objective usage metrics, assess learning or clinical-decision outcomes, and compare different clinical-support applications. Developing or adapting tools for rural primary care should also address Spanish-language search, offline access, ICD-10 codes, locally available medicines, referral pathways, and visual resources designed for rural clinical practice.

## DATA AVAILABILITY

The anonymized quantitative data and variable dictionary are available from the corresponding author upon reasonable request. Full interview transcripts are not publicly available because of the risk of participant identification in this small sample and the restrictions of the informed consent and ethics approval. The qualitative excerpts necessary to support the principal findings are included in the manuscript. Additional de-identified qualitative data may be available from the corresponding author upon reasonable request, subject to applicable ethical and consent restrictions.

## ACKNOWLEDGMENTS

The authors thank the participating physicians for sharing their time and experiences. We also thank the National University of San Marcos for facilitating contact with physicians eligible for invitation to the study.

## AUTHOR CONTRIBUTIONS

KDT: Methodology, investigation, formal analysis, data curation, writing—original draft, writing—review and editing.

PDS: Conceptualization, methodology, investigation, writing—review and editing.

SEA: Investigation, data curation, formal analysis, writing—review and editing.

LRM: Supervision, writing—review and editing.

All authors reviewed and approved the final version of the manuscript.

